## Supplementary material for "Rationale and protocol for a safety, tolerability and feasibility randomized, parallel group, double-blind, placebo-controlled, pilot study of a novel ketone ester targeting frailty via immunometabolic geroscience mechanisms": World Health Organization Trial Registration Dataset

**Supplemental information**

World Health Organization Trial Registration Dataset

| **Data category** | **Information**[**^32^**](https://www.spirit-statement.org/spirit-statement/references#32) |
| --- | --- |
| Primary registry and trial identifying number | ClinicalTrials.gov NCT05585762 |
| Date of registration in primary registry | 3 October 2022 |
| Secondary identifying numbers | NA |
| Source(s) of monetary or material support | Dr James B Johnson, philanthropic donation  Buck Institute Impact Circle, philanthropic donation.  BHB Therapeutics, provided active KE product free of charge. |
| Primary sponsor | Buck Institute for Research on Aging |
| Secondary sponsor(s) | NA |
| Contact for public queries | Brianna Stubbs, DPhil. |
| Contact for scientific queries | John Newman, MD, PhD |
| Public title | Buck Institute Ketone Ester RCT (BIKE) |
| Scientific title | A Randomized, Double-blind, Placebo-controlled, Parallel Group, Feasibility Pilot Study to Evaluate the Tolerability and Safety of a Novel Ketone Ester Ingredient in Healthy Older Men and Women. |
| Countries of recruitment | USA |
| Health condition(s) or problem(s) studied | Tolerance Safety Issues Aging |
| Intervention(s) | Ketone ester Non-ketone placebo (canola oil) |
| Key inclusion and exclusion criteria | Age >= 65 y  Stable health |
| Study type | Interventional Allocation: randomized Intervention model: parallel assignment Masking: double blind (subject, investigator, outcomes assessor) Primary purpose: basic science |
| Date of first enrolment | 31 January 2023 |
| Target sample size | 30 |
| Recruitment status | Recruiting |
| Primary outcome(s) | The primary outcome measure is the proportion of subjects reporting the same moderate to severe symptom (among dizziness, headache or nausea) occurring on more than one day within any given recall period (after week 0 - 2 acclimation period) when ketone esters are consumed daily for 12 weeks. |
| Key secondary outcomes | Change in safety labs from baseline to week 4 and week 12 in the incidence of abnormal laboratory test results.  Subjects consume a single serving of either 12.5 g or 25 g of ketone ester. After this, their short-term (regularly over 4h) blood ketone changes are measured using capillary blood sampling.  Exploratory geroscience biomarkers, physical and cognitive function, quality of life. |
